# Contamination rates in serially sampled sputum specimens obtained during tuberculosis treatment to capture culture conversion

**DOI:** 10.1101/2025.03.26.25324668

**Authors:** N Niemand, JA Rooney, S Malatesta, N Rawoot, TC Bouton, EJ Ragan, T Carney, LF White, M Farhat, CR Horsburgh, B Myers, RM Warren, KR Jacobson

## Abstract

Sputum cultures are the gold standard for tuberculosis (TB) diagnosis and treatment monitoring. However, cultures in MGIT liquid media are susceptible to microbial contamination, often rendering them uninterpretable. Research has shown that maintaining strict cold chains and supervised sample collection can reduce contamination rates, but few longitudinal studies with weekly sampling have explored this. Here we evaluated whether (1) the time between specimen collection and laboratory processing and (2) unsupervised specimen collection are associated with contamination rates. Additionally, we estimated contamination rates over the first 12 weeks of treatment and assessed clinical and behavioral predictors of contamination. We collected 3155 sputum specimens from 301 participants undergoing TB treatment. Contamination was lowest (12.3%) at treatment initiation, increased over the first few weeks, and stabilized around 30% from week 8 onwards. Samples collected without supervision were more likely to be contaminated at treatment initiation (p=0.048) and over the 12 weeks (p=0.028). We observed an inverse relationship between smear grade and contamination risk throughout the sampling period. These findings underscore the importance of supervised sputum collection to reduce contamination and provide ways to enhance the clinical and research value of weekly cultures, particularly those collected later in treatment. This is especially relevant for community-collected specimens used in monitoring treatment response.

## Introduction

Sputum culture remains the preferred method for *Mycobacterium tuberculosis* (Mtb) detection and drug susceptibility testing (DST).^1^ Culture’s sensitivity and usefulness in diagnostics and treatment response monitoring depend on sample quality.^2,3^ Culture contamination may lead to false negative or indeterminate results, increasing the burden on healthcare systems when repeated sample collection and testing are required and potentially delaying treatment initiation.^4–7^ Colony-forming units (CFUs) and time to positivity (TTP), measures associated with TB disease burden and level of infectiousness, and early bactericidal activity (EBA) for anti-TB medication assessments may be inaccurate and uninterpretable in contaminated samples.^8–10^ Additionally, contaminant DNA affects culture results and introduces false genetic variability in sequencing, including emerging methods for detecting drug resistance and strain diversity without the biohazard risks of traditional culture methods.^11^

Despite this, there is limited consensus on optimal collection, storage, transport, and handling practices to minimize contamination, and current protocols rarely assess sputum quality for TB diagnostics.^12,13^ The Stop TB Partnership and World Health Organization (WHO) recommend at least one specimen be collected in the early morning, with strict adherence to standardized laboratory protocols^14^, including refrigeration at 2-8 degrees Celsius (°C) within 1 hour of collection and processing within 48 hours.^15^ In practice, procedures are site- and/or laboratory-specific.^16,17^ As smoking cigarettes and eating can alter the lung and oral microbiome, an oral wash before collection is suggested to improve sample quality,^5,18–24^ as well as pooling specimens, various storage conditions, shortening transport intervals, and providing patient instruction and supervising collection, of which the latter has shown the most promise.^13,24–26^

The United States Center for Disease Control (CDC) proposes a 3-5% contamination target as an acceptable benchmark for a well-functioning biosafety level 3 (BSL-3) diagnostic facility.^27^ Achieving these standards in low-to middle-income country (LMIC) settings is often challenging due to structural and financial barriers. In LMICs, BSL-3 facilities are typically centralized in National Reference Laboratories or Tertiary Care Hospitals.^28^ Sputum specimens often require long-distance transport to reach these centralized locations, and maintaining the recommended cold chain is not always feasible. Information on how these delays impact contamination risk could identify regions where stricter protocols could improve diagnostic accuracy.

We aimed to evaluate rates and predictors of contamination in serially collected sputum specimens from individuals undergoing treatment for drug-susceptible pulmonary TB. Specifically, we examined the relationships between contamination, supervised specimen collection, and the time elapsed from sputum collection to MGIT inoculation during the first 12 weeks of treatment. Additionally, we assessed associations between sociodemographic, substance use, sputum quality, and clinical variables.

## Methods

### Study Setting and Participants

The Tuberculosis Treatment and Alcohol Use Study (TRUST) was a prospective longitudinal cohort that enrolled individuals initiating TB treatment to assess the impact of alcohol consumption on treatment response.^29^ Inclusion criteria were age ≥15 years, Mtb bacterial confirmation (positive GeneXpert MTB/RIF or Ultra and/or mycobacterial culture) without evidence of rifampicin resistance, not pregnant at enrollment, and living in Worcester, South Africa.^29^ Participants consented to an enrolment visit on the day of treatment initiation, daily Directly Observed Therapy (DOT) during the 6-month treatment period, and post-treatment visits up to 1 year after treatment completion. Participants were assigned a study DOT worker and followed during treatment, with sputum specimens collected weekly for 12 weeks post-treatment initiation and at the 5-month follow-up visit.^30^ For this analysis participants were excluded if no treatment initiation (week 1) sputum specimen was collected. Ethical approval was obtained from the Boston Medical Center Institutional Review Board (H-34970), the Health Research Ethics Committees from the South African Medical Research Council (EC011-5/2016), and Stellenbosch University (N17/04/039 RECIP_MRC_EC011-5/2016).

### Sputum Collection Procedures

Sputum specimens were considered collected under supervision if produced in the presence of a field team member (research nurse, trained staff, or DOT worker). Supervised collection involved the field team member guiding the collection, including mouth rinse, keeping the specimen jar closed until expectoration, and ensuring a good quality sputum (color, viscosity, and volume). Specimens were classified as unsupervised if participants produced them at home and then brought the sample to the research site (week 1 sample) or handed it to the DOT worker (daily visits at weeks 2–12). Participants were instructed to follow procedures used during supervised collections and produce sputum specimens first in the morning. However, we found that they often delayed collection until the DOT worker arrived (therefore resulting in supervised collection).

For the treatment initiation visit (referred to as week 1), specimens collected at the research site or delivered by the participant were immediately refrigerated at 2-8°C (Figure 1). Throughout weeks 2-12, DOT workers collected all sputum specimens on Tuesdays. Participants were asked to keep specimens cool until DOT workers collected them. Specimens were stored in cooler boxes with ice packs by the DOT workers until delivered to the research site for refrigeration. Sputum specimens were refrigerated at 2-8°C until twice-weekly transport (Wednesday and Friday), in a cooler box with ice packs, to the BSL-3 facility at Stellenbosch University, Cape Town (approximately one-hour drive), maintaining the cold chain and in compliance with WHO guidelines (Figure 1).^14^ No on-site decontamination was performed before transportation to the BSL-3 facility.

**Figure 1:**
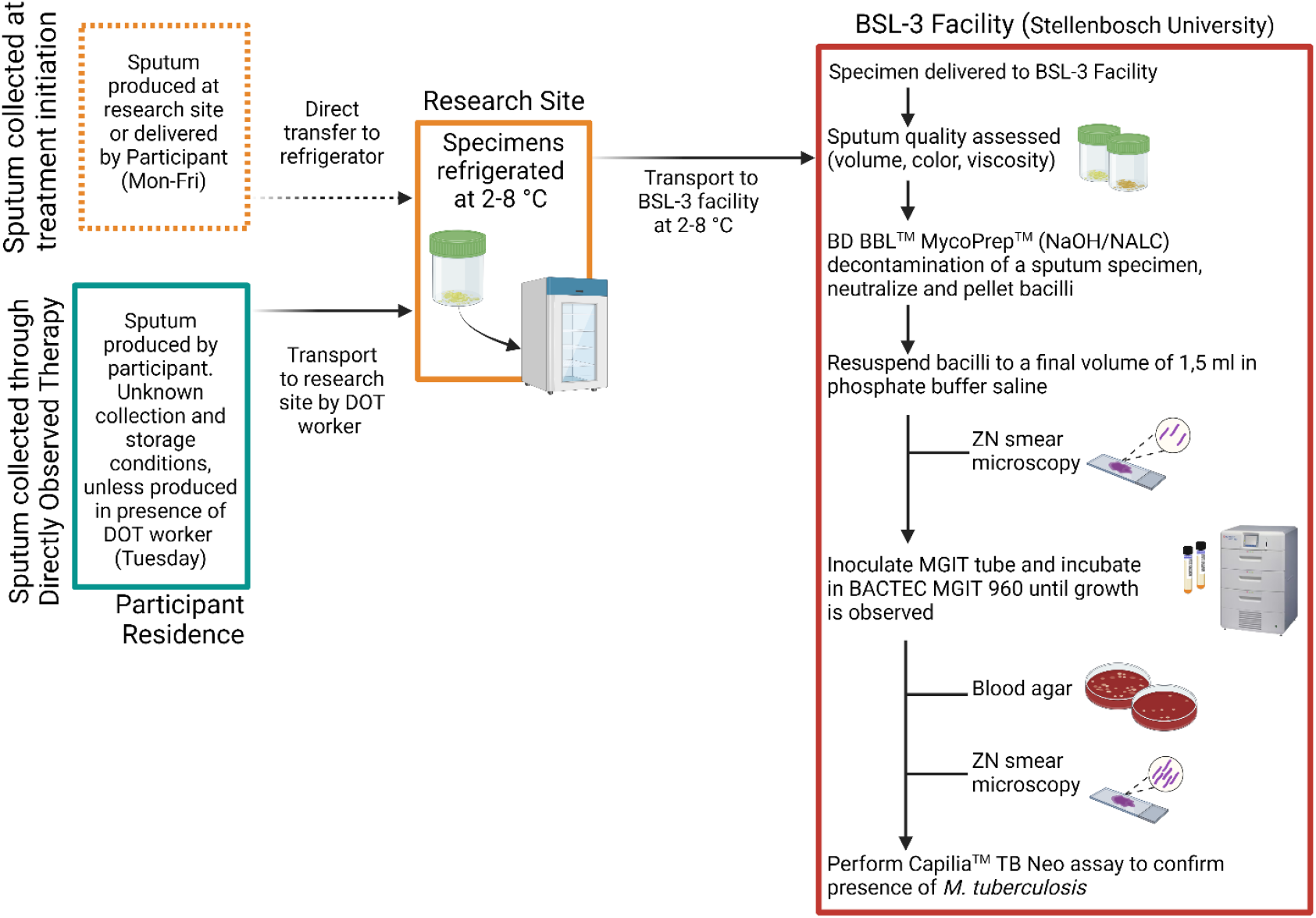
Sputum specimen collection and analysis strategy. Flow diagram illustrating the protocol for sputum specimen collection over the first 12 weeks of treatment initiation, interim storage conditions, and transport. The protocol followed within the BSL-3 facility for *M. tuberculosis* culture in Mycobacterial Growth Indicator Tubes for this study is outlined within the red rectangle. Created with BioRender.com

### Specimen Processing

Sputum quality (volume, color, viscosity) was recorded before decontamination with the BBL® MycoPrep™ Specimen Digestion/Decontamination Kit as per the manufacturer’s instructions (Figure 1). Briefly, sputum was processed by adding 2% sodium hydroxide (NaOH) with 0.5% N-acetyl-L-cysteine (NALC), with a final concentration of 1% NaOH. The specimen/NALC-NaOH mixtures were vortexed until a homogeneous sample was obtained, whereafter it was allowed to stand at ambient temperature for 15 min with occasional gentle inversion. Processing was completed by adding a phosphate buffer saline (PBS; pH 6.8) and centrifugation (15 min at 3000 x g). Concentrated pellets were resuspended to a volume of 1,5 ml in PBS. Ziehl-Neelsen (ZN) smear microscopic analysis was performed on all decontaminated crude sediments to evaluate and grade the presence of acid-fast bacilli (AFB). In addition, all sediments (0.5 ml) were inoculated into Mycobacteria Growth Indicator Tube 960 (MGIT, Becton Dickinson Microbiology Systems, USA) containing polymyxin B, amphotericin B, nalidixic acid, trimethoprim, and azlocillin (PANTA, BD, USA) and incubated in the BACTEC MGIT 960. Once a growth index of 70 growth units was observed, TTP was automatically recorded on the MGIT, and contamination was assessed by ZN smear microscopy and blood agar culture. All MGIT samples with a delayed TTP (longer than 20 days), with an AFB positive ZN smear were subjected to immune-chromatographic analysis, namely Capilia TB assay (TAUNS Laboratories, Inc), to confirm the presence of Mtb versus Non-Tuberculous Mycobacteria (NTM).

In this study, specimens were considered contaminated if a positive MGIT culture was observed to contain contaminant growth on either the culture ZN smear and/or blood agar (regardless of the presence of Mtb). This differs from the definition used within diagnostic laboratories, including the South African National Health Laboratory Services (NHLS), which considers specimens contaminated only if a positive MGIT culture was observed to contain growth, but no Mtb on the ZN smear (referred to as diagnostic definition).^31,32^

### Measures

At the treatment initiation visit, sociodemographic (age, race, biological sex, etc.) and substance use information was collected. Harmful alcohol use was defined as a phosphatidylethanol (PEth) blood test >49 ug/L and/or self-report with an Alcohol Use Disorders Identification Test (AUDIT) score >7.^33^ The Fagerström Test for Nicotine Dependence (FTND) was administered to screen for current tobacco use, while smoked drug use was defined by a positive urine drug test or self-reported use of methaqualone, methamphetamine, and/or cannabis.^34^ ZN smear is graded no AFB, scanty, 1+, 2+, and 3+.^35^

Chest radiographs captured lung cavitation while clinical data on HIV status, previous TB disease, and other comorbidities were extracted from medical charts. The treatment week variable was defined as the week since treatment initiation (week 1). Time to culture was defined as the interval in days between specimen collection and MGIT inoculation.

### Statistical Analysis

Contamination rates were calculated weekly as the percentage of contaminated specimens and stratified by AFB presence. The percentage of AFB-positive samples without contamination and the percentage of samples with no growth (Mtb nor contaminants) was calculated after 42 days.

Associations between contamination at treatment initiation and collection type (supervised versus unsupervised), time to culture (2 days or <2 days), sociodemographic factors, substance use, sputum quality, and clinical variables were examined using simple logistic regression models. Any variables associated with contamination (p-value <=0.1) were included in the multivariate analysis.

We fitted a multivariate logistic model with contamination as the outcome adjusting for age, sex, HIV status, smoked drug use, collection type (supervised vs. unsupervised), and time to culture. Although age, sex, and HIV status were not associated with contamination (p-value > 0.1), they were included in the model due to their potential impact on specimen quality.^36–41^

To examine longitudinal associations, we fitted a logistic regression model using Generalized Estimating Equations (GEE) to account for within-participant correlation of samples collected over time. We adjusted for age, sex, HIV status, and smoked drug use. Smear grade and collection type were included as time-varying covariates, with smear grade included to account for declining bacillary burden during treatment. We also adjusted for treatment week of the sputum collection. 0.6% of samples (N=15) were stored for ≥2 days before being processed, due to the low variability in this measure, it was not included in the longitudinal model.

All statistical tests were assessed using a 0.05 significance level. Analyses were conducted using R version 3.6.2.

## Results

Between May 2017 and May 2022, 3155 sputum specimens were collected from 301 participants who were diagnosed with pulmonary Mtb (positive GeneXpert MTB/RIF or Ultra, and/or mycobacterial culture). 301 samples were collected at treatment initiation visits and 2854 specimens across the remaining 11-week sampling period. At treatment initiation (week 1), 213 (70.8%) sputum specimens were smear-positive (grade scanty or above) and 274 (91.0%) were Mtb culture-positive. The median participant age was 38 years (IQR 27,48), 180 participants (59.8%) were male, and 85 (28.3%) were living with HIV (PLWH) (Table 1a).

**Table 1a:** Bivariate associations. Associations between sputum contamination in samples collected during treatment initiation visit (week 1 sputum specimen), and demographic, clinical, and collection condition variables. N=301

|  | median (IQR) or frequency (%) |  |  | Odds Ratio (95% CI) | p-value<sup>b</sup> |
| --- | --- | --- | --- | --- | --- |
|  | Total<br>n=301 | Contaminated<sup>a</sup><br>n = 37 | Non-contaminated<br>n = 264 |  |  |
| Male (sex) | 180 (59.8) | 18 (48.6) | 162 (61.4) | 0.60 (0.30, 1.19) | 0.143 |
| Age (years, ref. <30) |  |  |  |  | 0.990 |
| <30 | 87 (28.9) | 10 (27.0) | 77 (29.2) |  |  |
| 30-39 | 77 (25.6) | 10 (27.0) | 67 (25.4) | 1.15 (0.45, 2.96) |  |
| 40-49 | 73 (24.3) | 9 (24.3) | 64 (24.2) | 1.08 (0.41, 2.85) |  |
| >50 | 64 (21.3) | 8 (21.6) | 56 (21.2) | 1.10 (0.40, 2.96) |  |
| Living with HIV (n=300) | 85 (28.3) | 11 (29.7) | 74 (28.1) | 1.08 (0.49, 2.25) | 0.840 |
| Tobacco use | 205 (68.1) | 21 (56.8) | 184 (69.7) | 0.57 (0.28, 1.17) | 0.117 |
| Smoked drug use<sup>c</sup> | 165 (54.8) | 15 (40.5) | 150 (56.8) | 0.52 (0.25, 1.04) | 0.066 |
| Harmful alcohol use<sup>d</sup> | 186 (61.8) | 23 (62.2) | 163 (61.7) | 1.02 (0.51, 2.11) | 0.961 |
| Sputum volume (mL) (n=300) | 2 (1,4) | 3 (2,5) | 2 (1,4) | 1.13 (0.97, 1.29) | 0.093 |
| Sputum color (yes) (n=300) | 244 (81.3) | 28 (75.7) | 216 (82.1) | 0.68 (0.31, 1.61) | 0.348 |
| Supervised expectoration (n=276) | 263 (95.3) | 25 (86.2) | 238 (96.4) | 0.24 (0.07, 0.92) | 0.024 |
| Time to culturing ≥2 days<sup>e</sup> (n=300) | 142 (47.3) | 21 (56.8) | 121 (46.0) | 1.54 (0.77, 3.13) | 0.223 |
| Smear grade (ref. +++)<sup>f</sup> |  |  |  |  | 0.290 |
| +++ | 85 (28.2) | 10 (27.0) | 75 (28.4) |  |  |
| ++ | 48 (15.9) | 3 (8.1) | 45 (17.0) | 0.50 (0.06, 0.25) |  |
| + | 48 (15.9) | 10 (27.0) | 38 (14.4) | 1.97 (0.11, 1.74) |  |
| Scanty | 32 (10.6) | 3 (8.1) | 29 (11.0) | 0.76 (0.17, 2.75) |  |
| No AFB<sup>g</sup> | 88 (29.2) | 11 (29.7) | 77 (29.2) | 1.07 (0.43, 2.71) |  |
| Culture Positive<sup>h</sup> | 274 (91.0) | 33 (89.2) | 241 (91.3) | 0.79 (0.28, 2.81) | 0.676 |
| Cavitary TB disease<sup>i</sup> (n=294) | 191 (63.7) | 22 (59.5) | 169 (64.3) | 0.83 (0.41, 1.73) | 0.605 |

<sup>a</sup> Presence of contaminants on a blood agar plate or smear microscopy after detection of a positive MGIT culture
<sup>b</sup> All p-values represented were calculated using simple logistic regression models
<sup>c</sup> Defined as a positive urine drug test or self-reported use of methaqualone, methamphetamine, and/or cannabis
<sup>d</sup> Defined as a phosphatidylethanol (PEth) blood test >49 ug/L and/or an AUDIT score > 7
<sup>e</sup> Time from collection to processing and MGIT inoculation
<sup>f</sup> Observed with concentrated sputum smear microscopy
<sup>g</sup> Acid fast bacilli
<sup>h</sup> Presence of confirmed *M. tuberculosis*
<sup>i</sup> Lung cavitation identified using chest X-ray

**Table 1b:** Multivariate associations at treatment initiation. A multivariate logistic model, fit with contamination as the outcome and adjusting for time between sputum collection and processing, sex, age, smoked drug use, HIV status, and supervision of specimen collection. N=274

|  | OR<sup>a</sup> | 95% CI<sup>b</sup> | p-value |
| --- | --- | --- | --- |
| Supervised expectoration | 0.26 | 0.07, 1.10 | 0.048 |
| Time to culture, ≥ 2 days<sup>c</sup> | 1.58 | 0.71, 3.59 | 0.267 |
| Age (years, ref. <30) |  |  |  |
| 30-39 | 1.14 | 0.38, 3.55 | 0.813 |
| 40-49 | 0.84 | 0.25, 2.75 | 0.770 |
| >50 | 1.06 | 0.32, 3.50 | 0.923 |
| Smoked drug use | 0.48 | 0.20, 1.13 | 0.098 |
| Male (sex) | 1.07 | 0.46, 2.54 | 0.882 |
| HIV positive | 1.28 | 0.51, 3.00 | 0.587 |

<sup>a</sup> Odds Ratio
<sup>b</sup> Confidence interval
<sup>c</sup> Time from collection to processing and MGIT inoculation

### Rates of contamination over time

Contamination rates varied over the first 12 weeks of treatment, ranging from 12.3% (37/301) of week 1 (treatment initiation) samples to a maximum of 36.9% (85/232) of week 11 samples (Figure 3). When applying the diagnostic definition for contamination used within laboratories such as NHLS, contamination rates ranged from 1.3% (4/301) for week 1 samples to 31.7% (73/232) for week 11 samples (Figure 3).^32^ Of the 355 MGIT cultures with AFB and contamination according to ZN smear, 62 (17.5%) underwent confirmatory Capilia testing, resulting in 29 (46.8%) cases containing Mtb antigen (Figure 2).

**Figure 2:**
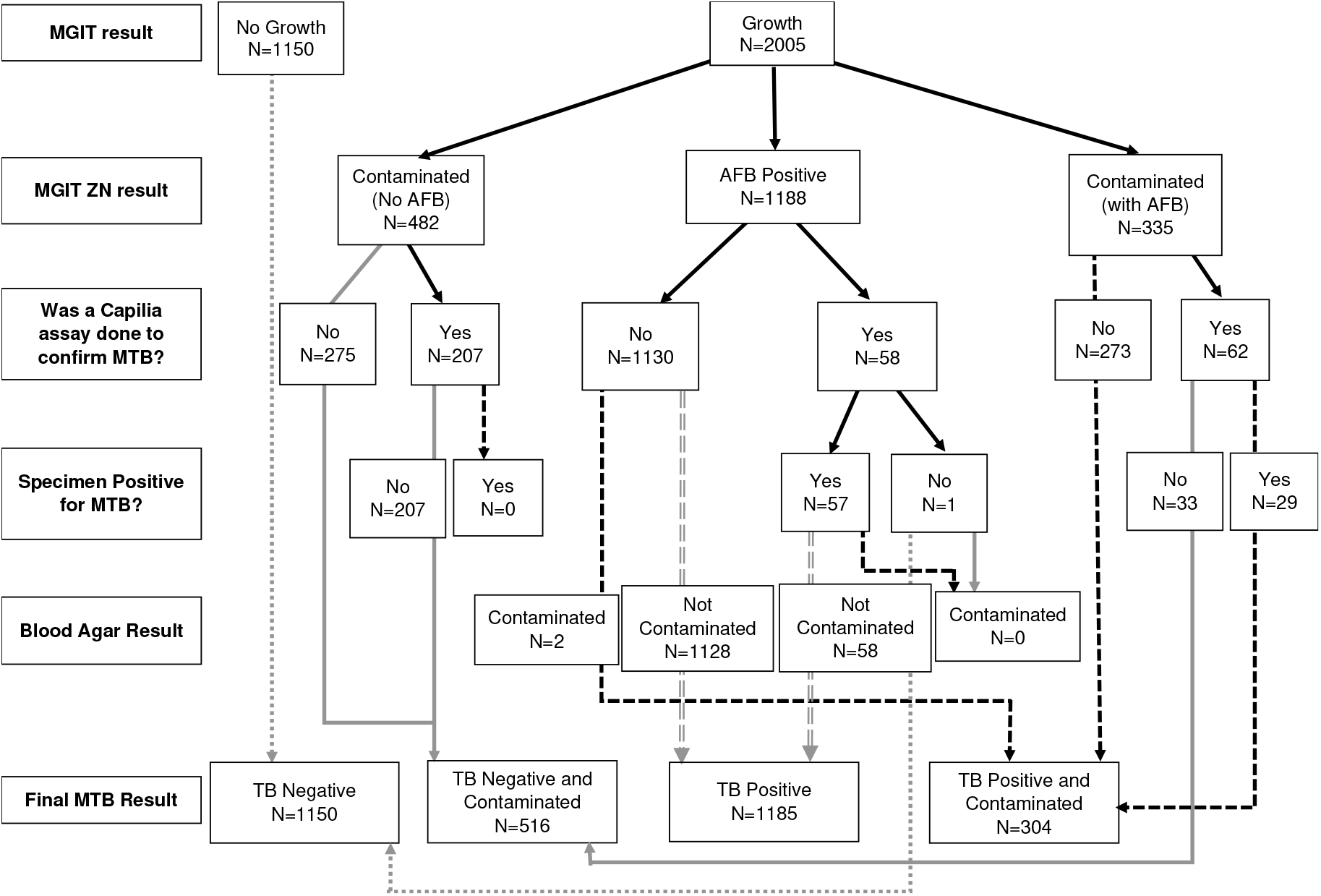
Classification of sample contamination. Flow diagram illustrating the classification of sample contamination after the BACTEC MGIT 960 system flags a culture positive for growth. If MGIT culture is not flagged as positive after 42 days of incubation in the BACTEC MGIT 960 system, it is regarded as culture negative (no growth). MGIT = Mycobacterial Growth Indicator Tubes; ZN = Zeihl Neelsen; AFB = acid-fast bacilli; Mtb = *M. tuberculosis*.

**Figure 3:**
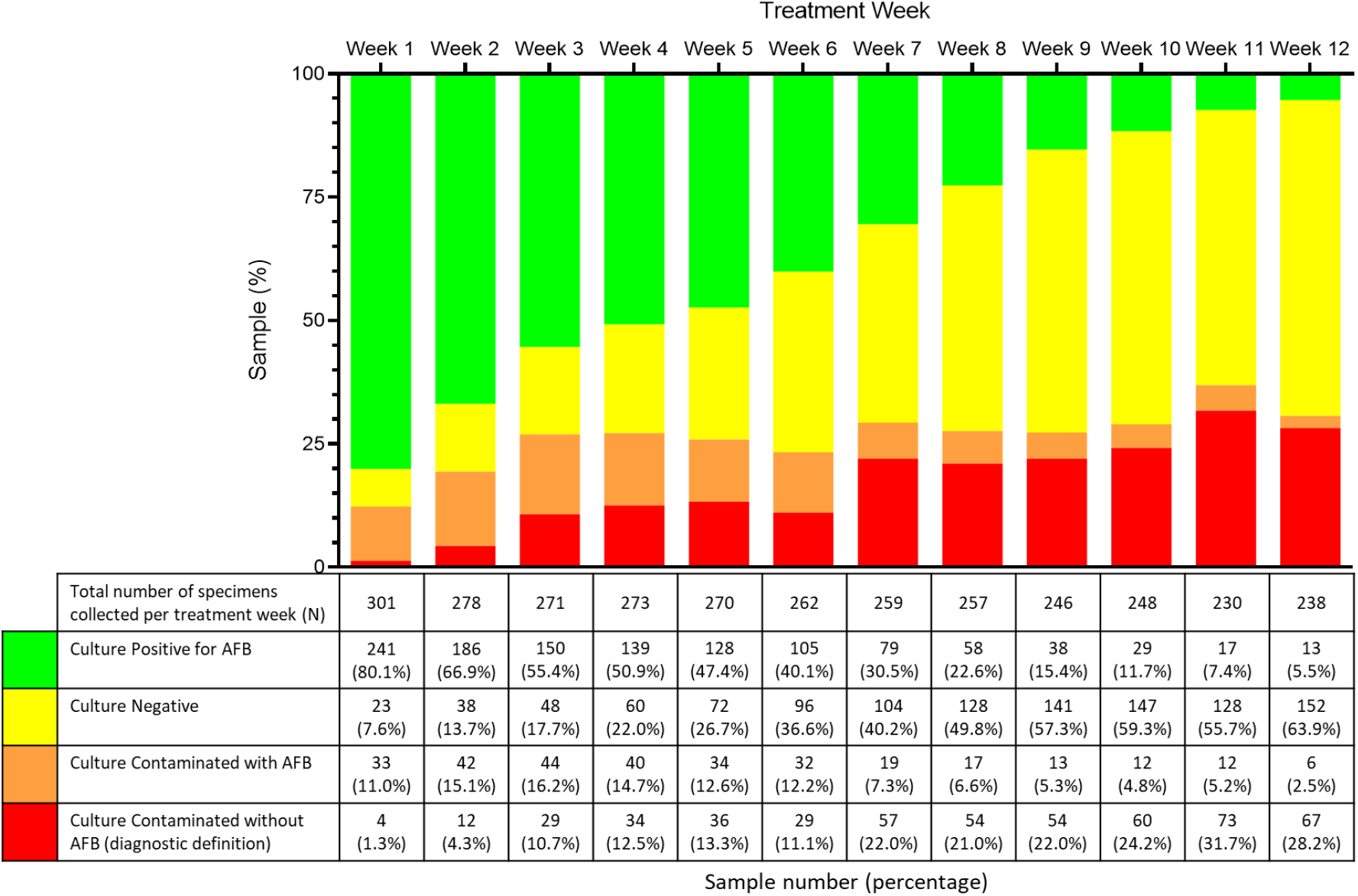
Contamination rates for 12 weeks following TB treatment initiation. N=3155. Green bars represent the percentage of AFB-positive samples with no contamination, and yellow bars indicate samples with no growth after 42 days. Orange bars indicate the percentage of samples contaminated in the presence of AFB. Red bars indicate the percentage of samples contaminated in the absence of AFB (diagnostic definition). The table indicates the total number of samples collected per treatment week, as well as the number and percentage of culture-positive or negative samples for the respective week following treatment initiation, with and without contamination.

### Predictors of contamination at treatment initiation

At treatment initiation supervised collection was associated with significantly less contamination (unadjusted OR=0.24, 95% CI 0.07-0.92, p=0.024) (Table 1a). Although not statistically significant, samples cultured 2 days or longer after collection (142/300) were more frequently contaminated (56.8% vs 46.0%, p=0.223).

The specimen color, cavitary TB disease, and culture positivity were not associated with contamination (Table 1a). Specimen volume, although marginally associated with contamination (p=0.093), was not included in the adjusted model due to concerns about its reliability. Harmful alcohol use and tobacco use were not associated with contamination rates, while the association between smoked drug use and contamination approached significance (OR 0.52, 95% CI 0.25-1.04, p=0.066) (Table 1a).

274 sputum samples (27 excluded due to missing covariate data) were included in the adjusted analyses of contamination at treatment initiation. Age, sex, HIV status, smoked drug use, and time to culture were not associated with contamination rates. There was a significant association of supervised collection with decreased contamination, with the odds of contamination consistent with unadjusted analysis (OR=0.26, 95% CI 0.07-1.10, p=0.048) (Table 1b).

### Predictors of contamination across the 12-week sampling period

The model evaluating predictors of contamination over time included 2915 sputum samples (Table 2); 218 samples were excluded due to missing covariate data. Over time, the supervised collection remained a significant predictor of reduced contamination (OR=0.79, 95% CI 0.64-0.98, p=0.031). A negative or scanty sputum smear grade was associated with a higher risk of contamination (OR=1.55, 95% CI 1.05-2.29, p=0.028 and OR=1.99, 95% CI 1.31-3.03, p=0.001; respectively) compared to a smear grade of 3+. Age, sex, HIV status, time to culture, and smoked drug use were not associated with contamination. A significant association was found between contamination and the treatment week of sputum collection, with each additional week increasing the odds of contamination (OR = 1.07, 95% CI 1.03–1.10, p < 0.001).

**Table 2:** Longitudinal associations across the 12 treatment weeks. A logistic regression model was fitted to account for the within-participant correlation of contamination status from treatment initiation up to treatment week 12. N=2915

|  | OR<sup>a</sup> | 95% CI<sup>b</sup> | p-value |
| --- | --- | --- | --- |
| Supervised expectoration | 0.79 | 0.64, 0.98 | 0.031 |
| Smear grade (ref. +++)<sup>c</sup> |  |  |  |
| No AFB<sup>d</sup> | 1.55 | 1.05, 2.29 | 0.028 |
| Scanty | 1.99 | 1.31, 3.03 | 0.001 |
| + | 1.39 | 0.93, 2.09 | 0.107 |
| ++ | 1.35 | 0.83, 2.20 | 0.223 |
| Age (years, ref. <30) |  |  |  |
| 30-39 | 0.97 | 0.71, 1.32 | 0.835 |
| 40-49 | 0.91 | 0.66, 1.25 | 0.553 |
| > 50 | 0.96 | 0.69, 1.34 | 0.804 |
| Smoked drug use | 1.06 | 0.83, 1.36 | 0.642 |
| Male (sex) | 1.02 | 0.80, 1.31 | 0.876 |
| Living with HIV | 1.14 | 0.90, 1.44 | 0.278 |
| Treatment Week<sup>e</sup> | 1.07 | 1.03, 1.10 | <0.001 |

<sup>a</sup> Odds Ratio
<sup>b</sup> Confidence Interval
<sup>c</sup> Observed with concentrated sputum smear microscopy
<sup>d</sup> Acid fast bacilli
<sup>e</sup> Timepoint at which sample was collected by field or DOTS workers across a 12-week sampling period following treatment initiation

## Discussion

Despite advances in sputum-based TB diagnostics, culture contamination remains a significant issue, leading to false negatives, indeterminate results, and unreliable analytic outcomes. Therefore, quality specimens are essential, and rigorous collection, storage, and transport protocols are required to ensure the optimal use of resources.

Our results indicate a large increase in culture contamination in our study from week 1 (12.3%) to week 2 (19.3%), likely due to change from clinic to field-based sampling with less supervision. Although specimen collection, transport, and processing procedures remained constant from week 2 onwards, contamination rates further increased in week 3 and remained relatively steady for the rest of the 12-week sampling period (23.3-36.9%). This aligns with previous research reporting contamination rates in TB diagnostic samples ranging from 0% at treatment initiation to 37% in follow-up samples.^25,26,42^ This is likely due to the reduced Mtb bacilli burden, allowing other microbes to grow while the sample processes for longer, and introduced oral and esophageal flora as sputum specimens become more difficult to produce in later treatment weeks. Our findings support this explanation, with increased odds of contamination among samples with a negative or scanty smear result compared to 3+ sputum smear microscopy grade and a significant association between the treatment week of specimen collection and contamination.

It is important to note that in the South African NHLS, contamination is defined as growth-positive MGIT culture with no AFB by ZN microscopy.^31,32^ Samples are only used for diagnostic purposes, as opposed to research laboratories, where downstream use of the Mtb isolates is essential and data is only interpretable if not contaminated. Our definition therefore considers a sample contaminated if other microbes are present on either ZN stain or blood agar, regardless of Mtb growth. Using the diagnostic laboratory definition, the contamination rate at treatment initiation in this study would be 1.3%, well below the proposed 3-5%.^27^

In 62 MGIT cultures with AFB and contamination that underwent confirmatory Capilia testing only 29 cases contained Mtb antigen – demonstrating the presence of non-tuberculous mycobacteria (NTM) frequently seen in patients receiving anti-TB treatment.^43^ Since NTM are ubiquitous environmental organisms, these NTM-positive cultures may indicate colonization or infection.^44^

Supervised sputum collection reduced contamination rates both at treatment initiation and in serial samples over the first 12 weeks of TB treatment. Previous reports have suggested contamination rates of 37% observed amongst samples collected at home without supervision.^26^ Supervised collection was associated with a 74% reduction in the odds of contamination for treatment initiation samples. In the longitudinal analysis, it was also linked to a 21% reduction in the odds of contamination, demonstrating its effectiveness in reducing contamination even in later-in-treatment samples. Contamination rates in weeks 8–12 ranged from 27.6% to 36.9%, but supervised collection, and proper instruction, likely helped mitigate contamination, particularly as the bacillary burden decreased over time.

At treatment initiation, samples inoculated for MGIT culture 2 days or longer after collection were observed to be slightly more frequently contaminated (56.8%). The lack of significance may reflect the small number of samples with delayed inoculation (47.3%). Our findings support that aiming for inoculation within two days of sample collection should yield reliable, low contamination rates, and are consistent with a South African study that found no significant association between contamination and time from collection to processing (0–4 days).^45,46^ However, previous studies have reported increased contamination with longer storage times.^47,48^ We were unable to assess the impact of delayed cold chain on contamination for samples collected during weeks 2–12, as there was little variability in the time from collection to culture. Maintaining a strict cold chain therefore seems crucial for samples collected later in treatment, when bacillary burden and sputum quality may decline.

No significant association was found with any substance use. In unadjusted analyses, smoked drug use appeared to have a protective effect; however, these results may be confounded by bacterial load, as higher bacterial load, more common among people who use drugs, may reduce the likelihood of contamination. This is supported by longitudinal models adjusted for smear grade, where the association was in the expected direction, although not significant.

Finally, the Stop TB Partnership and WHO guidelines specify that at least one specimen for Mtb testing be collected early morning to reduce contamination although in practice this may be challenging.^1^ One limitation is that DOT workers did not consistently capture when and under what conditions unsupervised specimen collections occurred. In addition, participants were asked to store specimens in a refrigerator after the morning expectoration, but since many participants did not have access to refrigerators, this level of detail was not captured.

## Conclusion

Measures like Mtb culture positivity, and analytic measures (e.g., TTP, DNA sequencing) can be inaccurate and uninterpretable when sputum cultures are contaminated. Our study benefits from serial sputum sampling over the first 12 weeks of TB treatment, allowing us to assess contamination rates while accounting for the decline in bacillary burden throughout treatment. This approach is novel, as other studies typically only report contamination data at baseline and treatment completion. Our findings emphasize the benefits of supervised sputum collection for TB diagnosis and monitoring treatment outcomes and show that while contamination rates generally increase in later-treatment samples, supervised collection can help mitigate this issue. These results can inform decentralized TB monitoring programs, emphasizing the importance of participant education and supervision during sample collection to enhance diagnostic accuracy and treatment effectiveness in a high-burden, LMIC setting.

## Data Availability

All data produced in the present study are available upon reasonable request to the authors

## Acknowledgments

The authors thank our study team, DOT workers, and study participants without whom our work would not be possible.

